# Downregulated Interferon Signalling in T Cells is Associated with Response to Vedolizumab in Inflammatory Bowel Disease

**DOI:** 10.64898/2026.05.11.26352882

**Authors:** Yasmine El Hajj, Rachael Slater, Christopher Probert, George Tang, Maria T. Abreu, Neha Mishra, Sofie Haglund, Stefan Schreiber, Ahmed N. Hegazy, Sven Almer, Phillip Rosenstiel, Paul A Lyons, Sreedhar Subramanian

## Abstract

**Background:** Vedolizumab, a gut-selective anti-integrin therapy, is effective in IBD, but response rates remain variable. Conventional clinical and biochemical markers, including C-reactive protein and faecal calprotectin, have limited predictive value. Although recent transcriptomic studies have implicated T-cell–related signatures in predicting vedolizumab response, these findings lack validation across independent cohorts.

**Methods:** We analyzed pre-treatment transcriptomic profiles from whole blood and T-cell subsets across five independent cohorts comprising 100 patients with UC and CD. The primary outcome was clinical response. Secondary outcomes included clinical and biochemical remission.

**Results:** Among the 100 patients, 61 were responders and 39 non-responders, with no significant baseline clinical differences. Gene set enrichment analyses revealed downregulation of interferon alpha and gamma signalling in responders’ baseline blood samples, a finding validated across independent cohorts. Downregulated interferon signalling at baseline was also observed in patients who achieved clinical and biochemical remission. To build a predictive model, an adaptive elastic net logistic regression model was applied to baseline whole-blood RNA-sequencing data. The classifier achieved an AUC of 1.0 in training, 0.71–0.83 in UC validation cohorts, and 0.64–1.0 in CD cohorts. Reduced interferon signalling was observed across CD4□ and CD8□ T-cell subsets, including regulatory T cells, suggesting a broad immune signature rather than cell-type specificity.

**Conclusions:** Downregulated interferon signalling in peripheral blood prior to treatment is a reproducible molecular signature predictive of vedolizumab response and biochemical remission. Whole-blood transcriptomics revealed a robust interferon-axis signal that predicted vedolizumab response across independent cohorts, with stronger performance in UC than CD. Given heterogeneous clinical endpoints and assessment windows, these data provide proof-of-concept that warrants validation with standardised, endoscopy-based outcomes.

## Introduction

An improved understanding of the pathogenesis of inflammatory bowel disease (IBD) has led to a rapid expansion in therapies that target intracellular signalling pathways, block key cytokines, or modulate immune cell trafficking to the gut. Vedolizumab, an anti-integrin therapy, blocks the interaction between α4β7 on lymphocytes and vascular addressin cell adhesion molecule 1 (MadCAM-1), thereby selectively preventing lymphocyte trafficking to the gut^1^. Vedolizumab was approved in 2014 following phase 3 clinical trials in both Crohn’s disease (CD)^2^ and ulcerative colitis (UC)^3^ which demonstrated its efficacy and safety. Response rates to induction therapy were variable, at 31% in CD^2^ and 47% in UC^3^ and, with similar variability observed in real-world cohorts^4–6^. This variability in response, combined with concerns of treatment cost and side-effects as well as the recent approval of several advanced therapies, highlight the need to identify response predictors to ensure that patients most likely to benefit receive effective therapy without delay.

There have been several attempts to identify clinical, biochemical and molecular predictors of response to vedolizumab. Clinical parameters such as disease severity^4, 5^, phenotype^4, 5^ and smoking^4, 5^ often show variable and inconsistent associations with response to vedolizumab, limiting their predictive ability. Even the recently developed vedolizumab clinical decision support tool (VDZ-CDST)^7^, which incorporates multiple clinical parameters and appears to be specific to vedolizumab, remains largely unadopted in clinical settings and only shows a modest correlation with response. The area under the curve (AUC) values for association with clinical remission range from 0.67 to 0.69 across the cohorts^8^. Commonly measured biochemical parameters such as C-reactive protein (CRP) and faecal calprotectin also exhibit only modest associations with response^4, 9^, thus providing limited utility as predictive biomarkers. Several additional parameters have been investigated as potential biomarkers of vedolizumab response, including faecal microbiota composition and function^10^; baseline α4β7 expression in the intestine^11^, and on peripheral blood T cells, B cells, and natural killer cells; along with saturation levels of α4β7-integrin on effector memory T cells^12, 13^.

Recent efforts have used transcriptomic profiling of whole peripheral blood^14^ and colonic biopsies^15^, alongside multi-modal approaches integrating CyTOF, flow cytometry, single cell RNA-sequencing, CITE-seq, BCR/TCR sequencing and proteomics^16^ to identify biomarkers of vedolizumab response and clarify its mechanism of action^17^. While response to vedolizumab has been associated with innate immune signatures such as macrophage polarisation^18^, two independent studies^19^ identified predictive signatures within T-cell subpopulations, aligning with vedolizumab’s mechanism of action. Abreu *et al* demonstrated that T regulatory (Treg) expressed genes were the best predictors of response in a mixed cohort of UC and CD patients^19^. Similarly, a prospective cohort study using multi-modal profiling identified enhanced predictive ability within CD4^+^ memory T cells (Tmem)^16^. However, these findings have not been validated across multiple independent cohorts and no clinically available biomarkers currently exist to predict response to vedolizumab.

To identify robust transcriptomic signatures and biomarkers predictive of vedolizumab response, we analysed pre-treatment RNA-sequencing data from whole peripheral blood and T-cell subsets extracted from peripheral blood across five independent cohorts of UC and CD vedolizumab-treated patients. Our findings identify a downregulated interferon signalling signature associated with subsequent vedolizumab response and demonstrate the feasibility of a blood-based gene expression classifier across multiple cohorts. This integrated approach advances the development of predictive biomarkers, paving the way for personalised treatment strategies in IBD.

## Methods

### Patient cohorts and samples

Five independent prospective cohorts were included in this study, comprising patients with either UC or both UC and CD (Table 1). Samples at baseline (n = 100) were collected from the five cohorts, and follow-up samples (n = 25) were aditionally collected from the Kiel (14 weeks post-treatment) and Stockholm (10-12 weeks post-treatment) cohorts. Bulk RNA-sequencing of peripheral whole blood was performed for the Liverpool, Kiel and Stockholm cohorts (Supplementary Figure 1). In addition, bulk RNA-sequencing of sorted T cell populations was available for the Liverpool and Miami cohorts. For the Berlin cohort, single-cell RNA-sequencing data were generated and subsequently aggregated into pseudobulk profiles for downstream analysis.

**Table 1.**
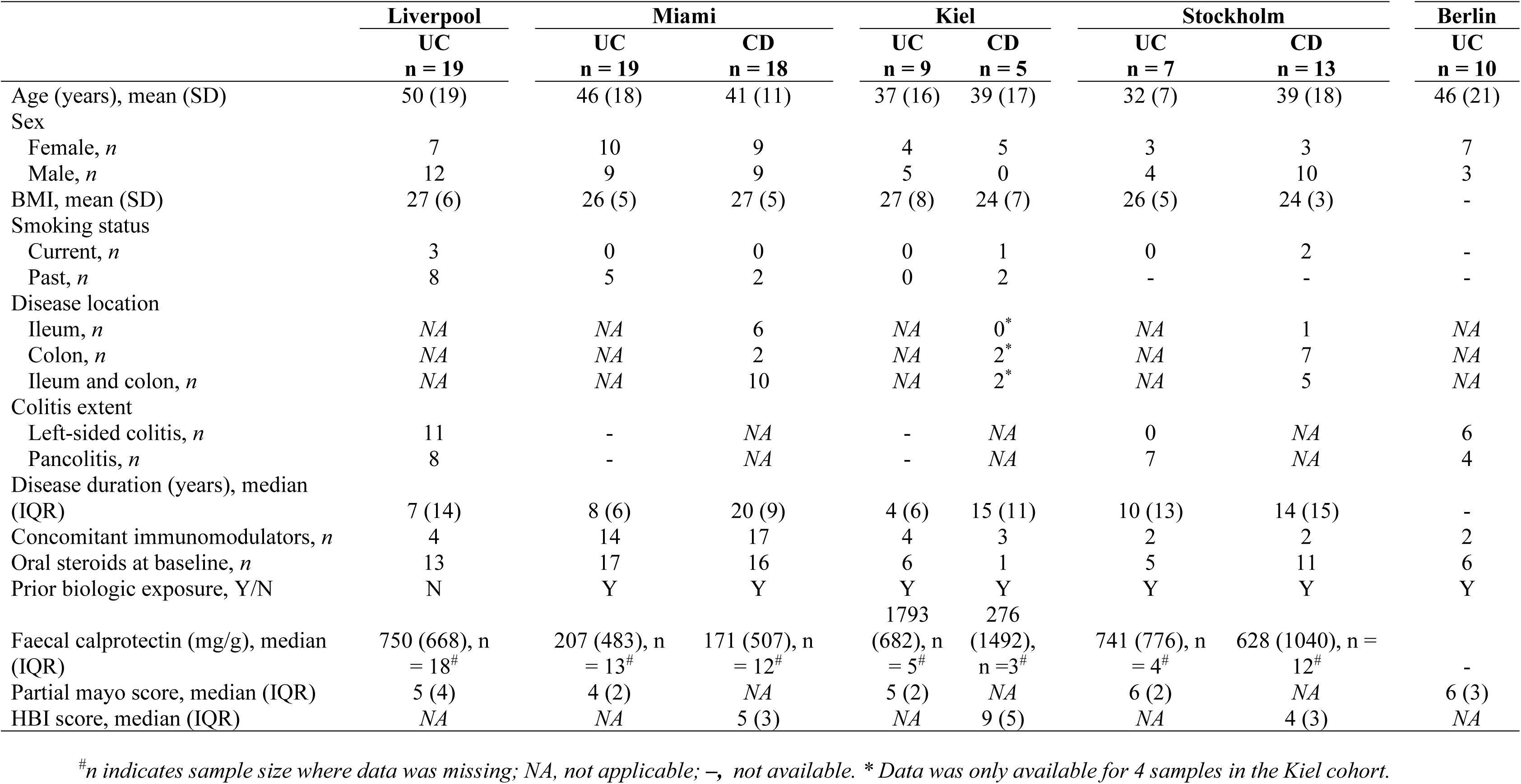
Baseline characteristics of patients across the 5 cohorts.

|  | Liverpool | Miami |  | Kiel |  | Stockholm |  | Berlin |
| --- | --- | --- | --- | --- | --- | --- | --- | --- |
|  | UC<br>n = 19 | UC<br>n = 19 | CD<br>n = 18 | UC<br>n = 9 | CD<br>n = 5 | UC<br>n = 7 | CD<br>n = 13 | UC<br>n = 10 |
| Age (years), mean (SD) | 50 (19) | 46 (18) | 41 (11) | 37 (16) | 39 (17) | 32 (7) | 39 (18) | 46 (21) |
| Sex |  |  |  |  |  |  |  |  |
| Female, *n* | 7 | 10 | 9 | 4 | 5 | 3 | 3 | 7 |
| Male, *n* | 12 | 9 | 9 | 5 | 0 | 4 | 10 | 3 |
| BMI, mean (SD) | 27 (6) | 26 (5) | 27 (5) | 27 (8) | 24 (7) | 26 (5) | 24 (3) | - |
| Smoking status |  |  |  |  |  |  |  |  |
| Current, *n* | 3 | 0 | 0 | 0 | 1 | 0 | 2 | - |
| Past, *n* | 8 | 5 | 2 | 0 | 2 | - | - | - |
| Disease location |  |  |  |  |  |  |  |  |
| Ileum, *n* | NA | NA | 6 | NA | 0* | NA | 1 | NA |
| Colon, *n* | NA | NA | 2 | NA | 2* | NA | 7 | NA |
| Ileum and colon, *n* | NA | NA | 10 | NA | 2* | NA | 5 | NA |
| Colitis extent |  |  |  |  |  |  |  |  |
| Left-sided colitis, *n* | 11 | - | NA | - | NA | 0 | NA | 6 |
| Pancolitis, *n* | 8 | - | NA | - | NA | 7 | NA | 4 |
| Disease duration (years), median (IQR) | 7 (14) | 8 (6) | 20 (9) | 4 (6) | 15 (11) | 10 (13) | 14 (15) | - |
| Concomitant immunomodulators, *n* | 4 | 14 | 17 | 4 | 3 | 2 | 2 | 2 |
| Oral steroids at baseline, *n* | 13 | 17 | 16 | 6 | 1 | 5 | 11 | 6 |
| Prior biologic exposure, Y/N | N | Y | Y | Y | Y | Y | Y | Y |
| Faecal calprotectin (mg/g), median (IQR) | 750 (668), n = 18<sup>#</sup> | 207 (483), n = 13<sup>#</sup> | 171 (507), n = 12<sup>#</sup> | 1793 (682), n = 5<sup>#</sup> | 276 (1492), n = 3<sup>#</sup> | 741 (776), n = 4<sup>#</sup> | 628 (1040), n = 12<sup>#</sup> | - |
| Partial mayo score, median (IQR) | 5 (4) | 4 (2) | NA | 5 (2) | NA | 6 (2) | NA | 6 (3) |
| HBI score, median (IQR) | NA | NA | 5 (3) | NA | 9 (5) | NA | 4 (3) | NA |

<sup>#</sup>*n* indicates sample size where data was missing; NA, not applicable; –, not available. \* Data was only available for 4 samples in the Kiel cohort.

#### Cohort 1 (Liverpool)

This study included a single-centre, prospective observational cohort consisting of 19 patients with active UC. All participants received vedolizumab therapy at baseline, and clinical response was evaluated at week 14. Prior to biological therapy, faecal samples were screened for infectious pathogens, including *Clostridium difficile*, as part of routine clinical care, and patients with concomitant infections were excluded. As patients in the Liverpool cohort were treatment-naïve prior to vedolizumab, this cohort was used as the discovery cohort.

#### Cohort 2 (Miami)

This was a prospective, single-centre cohort comprising patients with UC (n = 19) and CD (n = 18), as previously described^19^. Clinical response was evaluated at 14 to 22 weeks.

#### Cohort 3 (Kiel)

This prospective cohort comprised 9 patients with UC and 5 patients with CD, as previously described^20^. All patients had clinically active disease confirmed by clinical indices, endoscopy and laboratory assessments prior to induction therapy. Response to therapy was evaluated at week 14.

#### Cohort 4 (Stockholm)

This single-centre cohort included 7 patients with UC and 13 with CD who had previously failed at least one anti-TNF-α therapy, as described in Haglund et al^14^. Treatment response was assessed between weeks 10 and 12.

#### Cohort 5 (Berlin)

This single-centre cohort included 10 patients with UC, as previously described^16^. Treatment response was assessed at week 30.

### Treatment

All patients received vedolizumab at a fixed induction dosing of 300 mg at weeks 0, 2 and 6. Additional dosing at week 10 was at the discretion of the treating physician

### Clinical response, clinical and biochemincal remission definitions

A pragmatic clinical response definition (≥25% reduction in partial Mayo or HBI between 12–22 weeks) was selected to maximize cross-cohort inclusion. Heterogeneity in assessment windows is expected to bias associations toward the null; therefore, the observed cross-cohort replication likely underestimates the true effect. More stringent endpoints, such as endoscopic response and remission, may better reflect mucosal healing, however these measures were not uniformly available across cohorts and would have substantially reduced sample size. Secondary outcomes included (i) clinical remission, defined as a partial Mayo ≤ 2 at week 14 among UC patients with a baseline partial Mayo > 2^21^ and (ii) biochemical remission, defined as fecal calprotectin <250 µg/g at week 14 in patients with a baseline value >250 µg/g at initiation of therapy^22^. Evaluation of the secondary outcomes was restricted to the Liverpool cohort due to data availability.

### RNA isolation, sequencing and microarray analysis

#### Liverpool cohort

At enrolment, 10 mL of blood was collected in citrate tubes and 2.5 mL was collected into PAXgene® Blood RNA Tubes (BD Biosciences) in the morning to minimise circadian variation in gene expression and processed immediately^23^. Peripheral blood mononuclear cells (PBMCs) were isolated by density centrifugation and CD4^+^ and CD8^+^ T cells were positively selected using magnetic beads as previously described^24^. Total RNA was extracted from whole blood using the PreAnalytiX® PAXgene® Blood RNA Kit (IVD) (Qiagen), and from CD4^+^ and CD8^+^ T cells using the RNeasy Mini Kit (Qiagen) following the manufacturer’s instructions. RNA quality was evaluated with an Agilent Bioanalyser 2100, and RNA quantity was measured using a NanoDrop ND-1000 spectrophotometer.

RNA-sequencing libraries from whole blood samples were prepared using the NEBNext Ultra II Directional RNA Library Prep Kit (New England Biolabs). Sequencing was performed on a NovaSeq 6000 instrument (Illumina). Raw sequencing reads were processed as previously described^25^ and the reads were aligned to the human reference genome (build 38), a matrix of gene counts was generated from the resulting BAM files.

Total RNA isolated from CD4^+^ and CD8^+^ T cells was converted to cDNA and hybridised to the GeneChip Human Genome 2.0 ST array. Arrays were scanned using a GeneChip Scanner 3000. Raw data (.CEL files) were processed in R using the robust multi-array average method^26^. To account for batch effects, the ComBat function from the Surrogate Variable Analysis package was employed^27^ then the normalised data was analysed in R.

#### Miami, Kiel, Stockholm and Berlin cohorts

The full methodology for sample collection, library preparation and sequencing for the Miami, Kiel, Stockholm and Berlin cohorts have been described in detail elsewhere^14, 16, 19, 20^. For the Miami cohort, Tmem and Treg cells were sorted from PBMCs, followed by RNA extraction and sequencing^19^. For the Stockholm cohort, raw sequencing reads were processed as previously described^25^ and the reads were aligned to the human reference genome (build 38), then a matrix of gene counts was generated from the resulting BAM files. For the Kiel cohort, single-cell RNA-sequencing data was processed using the Seurat^28^ pipeline in R, mapped to a CITE-seq reference^29^, and subsequently aggregated into pseudobulk profiles.

The gene expression data for the Miami, Kiel and Berlin cohorts are available in the Gene Expression Omnibus database under accession numbers GSE184593, GSE191328 and GSE261334 respectively. Metadata and RNA-sequencing data for the Stockholm cohort are available from the investigators upon request.

### Statistical analysis

Continuous normally distributed variables have been summarised as means (sd, standard deviation), non-normally distributed continuous varibales as medians (interquitile range, IQR). We conducted univariate analysis using t-tests, Mann-Whitney U test and Chi-square test for continuous parametric, continuous non-parametric and categorical data, respectively. Where counts were small, Fisher’s Exact test was used to compare categorical data. Multiple hypothesis testing was accounted for using the Bonferroni correction.

### Differential expression analysis

Differential gene expression between responders and non-responders to vedolizumab was conducted in R using DESeq2^30^ for whole blood and sorted bulk RNA-sequencing datasets, as well as for pseudo-bulk RNA-sequencing data derived from the single-cell RNA-sequencing experiment, and using LIMMA^31^ for microarray data. When necessary, Ensembl gene IDs were mapped to gene symbols using the BioMart “hsapiens_gene_ensembl” dataset. Significance thresholds were set at an adjusted p-value of <0.05 and an absolute log_2_ fold change of ≥1. Differential expression results were visualised and interpreted based on these criteria. Gene set enrichment analysis (GSEA) was performed using the fgseaMultilevel algorithm^32^. Genes were pre-ranked based on test statistics derived from the differential expression analysis. Enrichment was tested against three gene set collections downloaded from the Molecular Signatures Database (MSigDB): (1) Hallmark gene sets (50 sets), (2) C2 canonical pathways (3,795 sets) and (3) C5 gene ontology biological processes (7,608 sets). Pathways were tested using a parametric t-test, with significance determined by an adjusted p-value <0.05. Additionally, specific gene signatures for CD4^+^ and CD8^+^ T cell exhaustion, derived from published studies^33, 34^, were included in enrichment analyses to evaluate immune exhaustion pathways related to treatment response.

### Predictive model building for early vedolizumab response

To develop a predictive model of early response to vedolizumab, a binomial regression with an adaptive elastic-net penalty was applied to variance-stabilised whole blood RNA-sequencing count datafrom the Liverpool cohort. The adaptive elastic net was selected for its ability to handle high-dimensional data, address multicollinearity and produce sparse, interpretable models by combining ridge and lasso penalties^35^. Model training was performed using the glmnet package in R, with 3-fold cross-validation and a maximum of 1,000 iterations. The optimal regularisation parameter (lambda.min) was used for prediction. To mitigate class imbalance between responders and non-responders, inverse frequency weights were assigned to each sample, with weights proportional to the inverse of class sample size and normalised to sum to one.

### Data availability

Research materials supporting this publication are not publicly available but may be accessed by contacting the corresponding authors.

## Results

A total of 100 patients treated with vedolizumab across five cohorts were included in the final analysis: 19 from Liverpool, 37 from Miami, 14 from Kiel, 20 from Stockholm and 10 from Berlin. The baseline characteristics of these subjects are summarised in Table 1. Overall, 61 patients were classified as responders and 39 as non-responders after induction therapy. Response rates by cohort were as follows: Liverpool cohort (n = 15, 79%), Miami cohort (n = 20, 54%), Kiel cohort (n = 10, 71%), Stockholm cohort (n = 10, 50%), and Berlin cohort (n = 6, 60%).

### Relationship between clinical factors and response

Where data was available (Liverpool, Miami, Kiel and Stocklhom cohorts), we compared baseline clinical and biochemical variables between responders and non-responders (Supplementary table 1). There were no significant differences in any of the variables examined including prior biologic exposure.

### Identification of an interferon response signature in non-responders

To explore potential pre-treatment molecular predictors of vedolizumab response, we performed a transcriptomic analysis of baseline whole blood samples from the Liverpool cohort. The goal was to identify genes or pathways whose expression levels could distinguish patients who would later respond or fail to respond to therapy. We compared the baseline gene expression profiles of patients who achieved clinical response at week 14 with those who did not (Supplementary table 2). This analysis revealed only two genes (*RAP1GAP* and *ANKRD29*) that were significantly differentially expressed between responders and non-responders (Figure 1A). This finding is broadly in keeping with Haglund et al.^14^, who reported no individual gene differences in whole blood between response groups, suggesting that pathway-level analysis might reveal more robust predictors.

**Figure 1.**
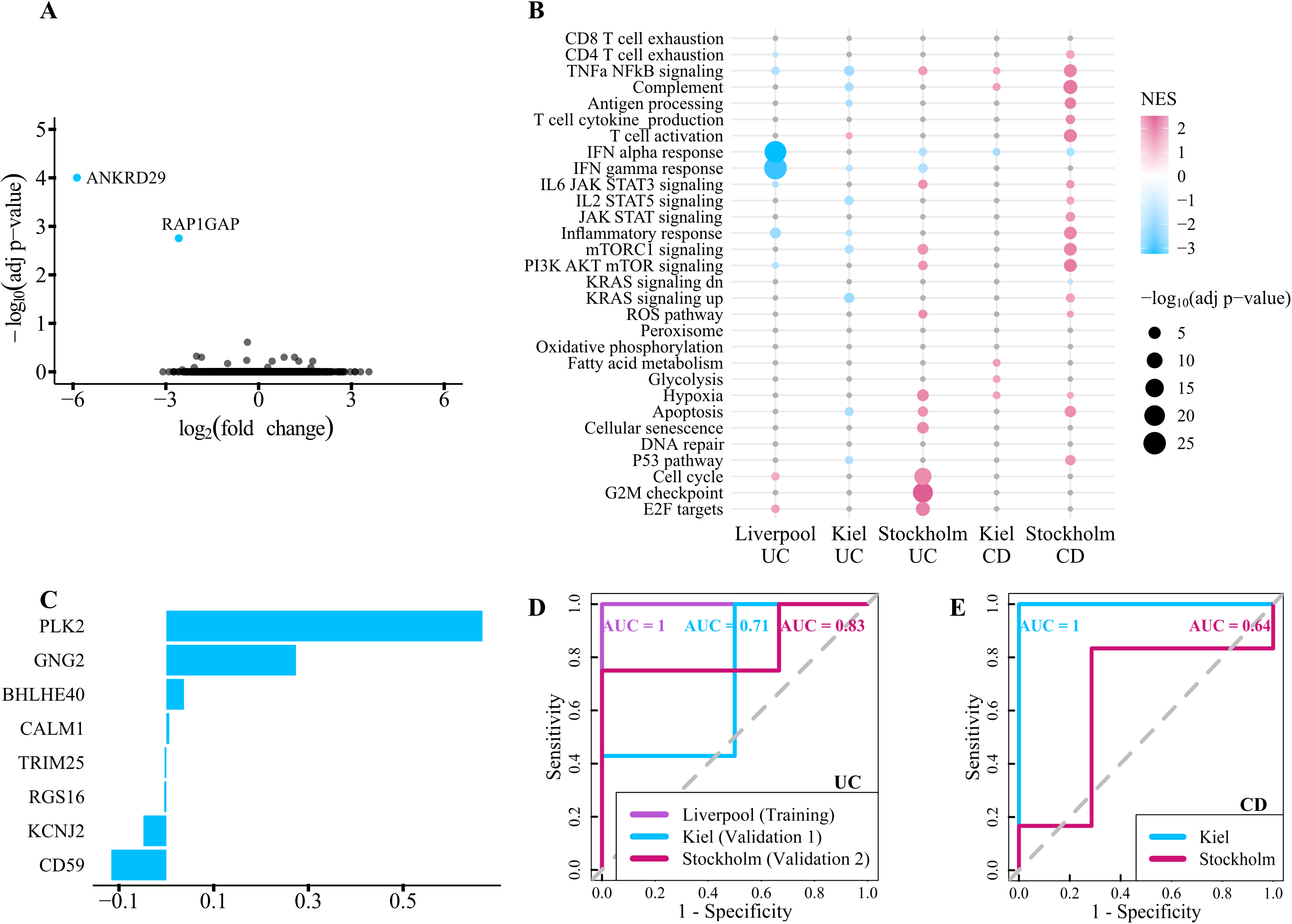
Transcriptomic predictors of vedolizumab response in whole blood across the Liverpool, Kiel and Stockholm cohorts. (A) Volcano plot of differentially expressed genes between vedolizumab responders and non-responders at baseline in whole blood from the Liverpool cohort. The two genes with |fold change|□>□1 and adjusted p-value□<□0.05 are shown in blue. (B) Pathways enriched at baseline in the whole blood transcriptomes of responders vs. non-responders across the Liverpool, Kiel and Stockholm cohorts. Blue and pink dots represent pathways significantly down or upregulated in responders, respectively. (C) Bar plots showing the top biomarkers and their weights in the whole blood-based computational model for predicting vedolizumab response. Receiver operating characteristic (ROC) curves and area under the curve (AUC) values for vedolizumab response prediction in the Liverpool, Kiel and Stockholm cohorts across (D) UC and (E) CD patients.

Pathway analysis using gene set enrichment analysis (GSEA) uncovered a broader transcriptional pattern: responders exhibited significant downregulation of inflammatory pathways, particularly interferon alpha (Padj = 1×10^−23^) and interferon gamma signalling (Padj = 3×10^−26^) (Figure 1B). These interferon pathways were also significantly downregulated in responders at baseline in independent cohorts of UC patients from Kiel (Padj = 0.03 for interferon gamma) and Stockholm (Padj = 0.01 for interferon alpha, Padj = 0.002 for interferon gamma), as well as CD patients from Kiel (Padj = 0.02 for interferon alpha) and Stockholm (Padj = 9×10^−3^ for interferon alpha) (Figure 1B). These findings suggest that suppressed interferon responses in whole blood prior to treatment may serve as a biomarker predictive of response to vedolizumab.

### Development and validation of a whole blood transcriptomic biomarker for early vedolizumab response

While pathway enrichment analyses provide valuable biological insights, they are not directly translatable to routine clinical practice. To overcome this limitation, we developed a predictive biomarker model utilising baseline whole blood RNA-sequencing count data. Using the Liverpool cohort as the training set, we applied a classic elastic net regression approach, optimising the alpha parameter to 0.5 to balance penalisation between L1 and L2 norms. This model identified eight key genes with the highest predictive weights (Figure 1C). In the training cohort, the elastic net classifier achieved perfect discrimination, with all performance metrics, sensitivity, specificity, precision, accuracy and area under the receiver operating characteristic curve (AUC), equal to 1 (Figure 1D, Table 2).

**Table 2.** Confusion matrix: model performance in predicting responders and non-responders relative to observed outcomes in the Liverpool, Kiel and Stockholm cohorts.

|  | **Liverpool (training)** |  | **Kiel and Stockholm (validation)** |  |  |  |
| --- | --- | --- | --- | --- | --- | --- |
|  |  |  | *Observed outcome* |  |  |  |
| ***Prediction*** | UC<br>Responders | UC Non-responders | UC<br>Responders | UC Non-responders | CD<br>Responders | CD Non-responders |
| Responders | 14 | 0 | 10 | 1 | 4 | 5 |
| Non-responders | 0 | 4 | 1 | 4 | 5 | 4 |

We then validated the predictive model in the two independent cohorts where whole blood baseline RNA-sequencing count data was available (Kiel and Stockholm), which included patients with both UC and CD. In UC, external validation yielded AUCs of 0.71 and 0.83 across centres (Figure 1D). In these UC validation cohorts, the sensitivity was 91%, the specificity was 80% and the overall accuracy was 88% (Table 2). Model performance in CD was variable (AUC 0.64–1), warranting CD-focused validation and potential disease-specific modeling (Figure 1E).

### Clinical and biochemical remission

11 out of 16 patients in the Liverpool cohort achieved clinical remission, while 13 out of 17 achieved biochemical remission. The downregulated interferon-associated transcriptional signal at baseline was also observed in patients achieving clinical remission (Padj = 2×10^−14^ for interferon alpha, Padj = 9×10^−16^ for interferon gamma) and biochemical remission (Padj = 1×10^−25^ for interferon alpha, Padj = 3×10^−39^ for interferon gamma). In addition, the elastic net model retained good predictive performance, achieving an AUC of 0.75 for clinical remission and 0.88 for biochemical remission. These findings support the biological relevance of the primary response definition used across cohorts.

### Identification of cellular sources underlying the interferon signature

To determine the specific cellular contributors to the observed interferon-alpha and –gamma signatures associated with response to vedolizumab, we analysed gene expression data from both sorted T cell populations and single-cell RNA-sequencing of peripheral blood mononuclear cells.

Initially, baseline GSEA was performed on transcriptomic data from magnetically sorted CD4□ and CD8□ T cells obtained from the Liverpool cohort. Comparing responders and non-responders, we observed significant downregulation of the interferon-alpha (Padj = 2×10^−23^ in CD4□ and Padj = 1×10^−20^ in CD8□ T cells) and interferon gamma signatures (Padj = 1×10^−27^ in CD4□ and Padj = 2×10^−22^ in CD8□ T cells) in responders across both subsets (Figure 2A). To further validate and refine these findings, we extended the analysis to the Miami cohort, performing GSEA on blood-derived Treg and Tmem cells. The interferon signatures were significantly reduced only within the Treg subset in UC (Padj = 1×10^−12^ for interferon alpha, Padj = 1×10^−8^ for interferon gamma) and CD cohorts (Padj = 9×10^−5^ for interferon gamma), consistent with prior reports that Treg gene expression signals are more predictive of response in this cohort^16^ (Figure 2A).

**Figure 2.**
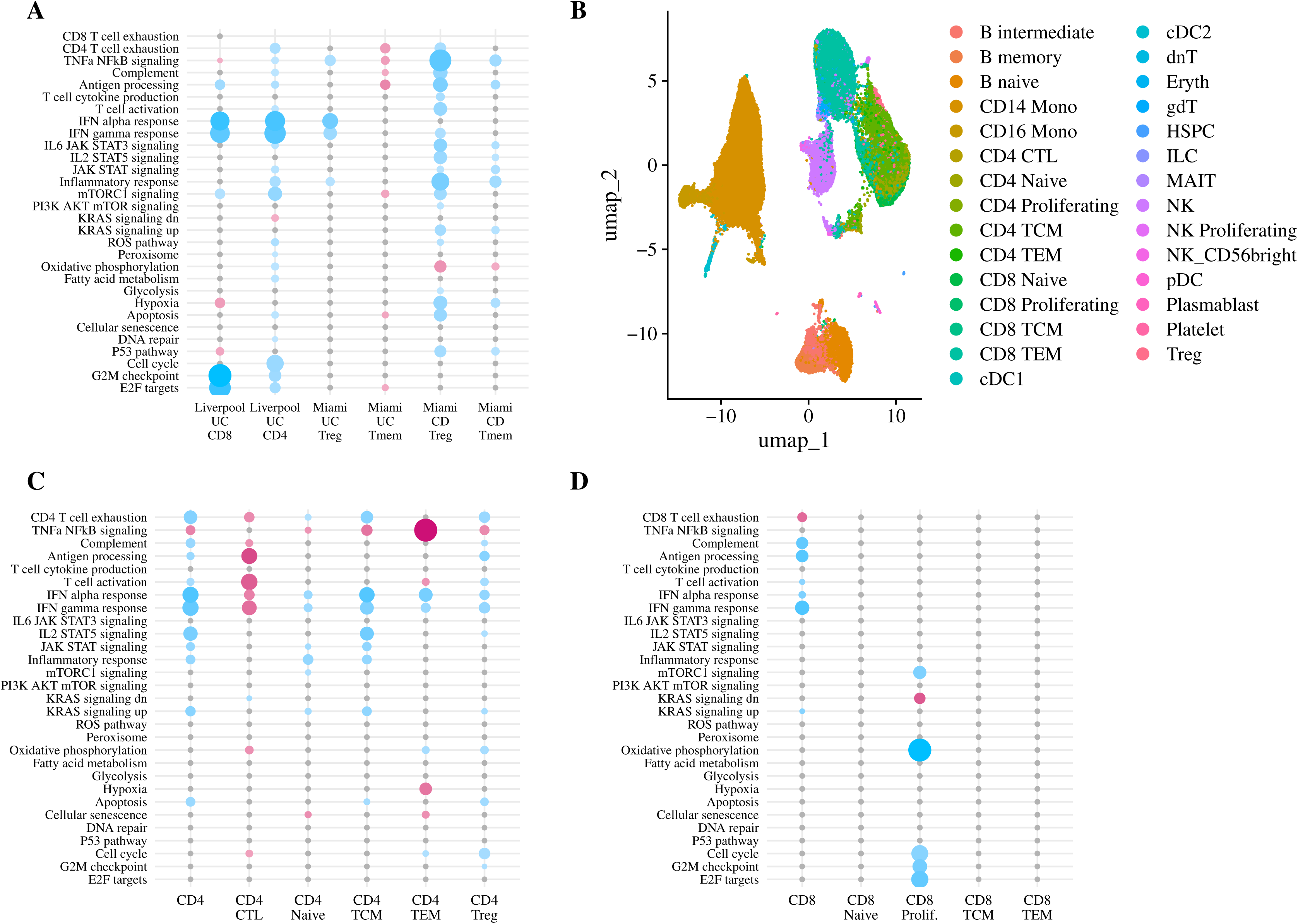
Transcriptomic predictors of vedolizumab response in T cell subsets across the Liverpool, Miami and Berlin cohorts. (A) Dot plot showing enriched pathways at baseline in responders versus non-responders in CD4^+^ and CD8^+^ T cells from the Liverpool cohort, and in T memory (Tmem) and T regulatory (Treg) cells from the Miami cohort. (B) UMAP of PBMCs from single-cell RNA-sequencing analysis in the Berlin cohort. (C,D) Dot plot showing enriched pathways at baseline in responders versus non-responders in (C) CD4^+^ and (D) CD8^+^ T cell subsets from the Berlin cohort.

Next, to achieve a higher-resolution understanding of the cellular sources, we analysed baseline peripheral blood single-cell RNA-sequencing data from the Berlin cohort. The results echoed the bulk analyses, showing a significant downregulation of interferon alpha and gamma in both CD4□ (Padj = 7×10^−7^ for interferon alpha, Padj = 6×10^−7^ for interferon gamma) and CD8□ T cells (Padj = 0.01 for interferon alpha, Padj = 1×10^−9^ for interferon gamma) of responders compared to non-responders (Figures 2B-D). The limited number of samples (n = 10) may have reduced power to identify specific CD8□ T cell subsets contributing to the interferon signal. Within the CD4□ T cell compartment, multiple subsets, including central memory (Padj = 3×10^−6^ for interferon alpha, Padj = 6×10^−5^ for interferon gamma), effector memory (Padj = 5×10^−5^ and 7×10^−3^), naïve (Padj = 2×10^−2^ for both signatures) and regulatory T cells (Padj = 5×10^−3^ and 2×10^−3^), contributed to the interferon responses. These findings suggest that the interferon signature in treatment response is neither restricted to a single T cell subset nor limited to either CD4□ or CD8□ lineages, but involves multiple T cell populations across both lineages.

### Characterisation of interferon signalling post-treatment

To determine if the interferon signatures remain suppressed post-treatment, we analysed interferon alpha and gamma inducible genes in responders from the Kiel (14 weeks) and Stockholm (10–12 weeks) cohorts. Interferon signatures were consistently downregulated across all post-treatment responders (Supplementary Figure 2), in line with findings from Haglund et al.^14^, confirming that reduced interferon signalling is a robust marker of therapeutic response.

## Discussion

Our study provides evidence that pre-treatment transcriptomic profiles, particularly the interferon alpha and gamma signatures, are associated with response to vedolizumab in patients with UC and CD. At baseline, responders exhibit significantly lower interferon pathway activity in peripheral blood relative to non-responders. The inclusion of 100 patients across five distinct cohorts allowed us to cross-validate this observation in diverse contexts. Because the interferon-associated signal remained stable across different sequencing approaches and clinical backgrounds, the findings are unlikely to be simply a product of overfitting. Rather, this cross-cohort consistency demonstrates the broad generalisability of the signature. While the discovery cohort was limited to treatment-naïve patients, the replication of these associations in independent cohorts, including those with prior biological therapy, confirms that the transcriptomic signal is independent of treatment history.

We developed an elastic net predictive model using baseline whole blood transcriptomics. While trained on a treatment-naïve cohort, the model’s high accuracy was sustained during independent validation, demonstrating that the signature is robust and not overfitted to the initial training set. Despite being trained exclusively on UC samples, the model retained predictive ability in CD, albeit with more variable performance, in keeping with the presence of shared transcriptomic features of vedolizumab response across disease subtypes. This suggests that a parsimonious gene signature could be practically applied to stratify patients before commencing therapy, facilitating personalised treatment decisions and potentially avoiding ineffective therapies.

Differential expression of interferons aligns with IBD pathophysiology and previous studies which identified interferon genes as predictors of biological therapy response. Numerous IBD-associated genetic variants are linked to type I interferon signalling pathway, including SNPs linked to *IFNGR2*, *IFNAR1*, *JAK2*, *TYK2*, *STAT1*, *STAT3*, *MDA5* and *IRF4*^36, 37^. Interferon-gamma, in particular, affects gut barrier and vasculature, contributing to IBD pathogenesis^38^. It also promotes CD4^+^ T cell migration across the endothelial monolayer^39^, which may lead to inflammation in the lamina propria and a lack of response to vedolizumab. Increased interferon signalling in non-responders may also reflect expansion of specific immune populations, including proliferating CD4□ memory and Th1 cells^16, 40^. Changes in type I interferon gene expression are linked to responses to biological treatments in immune-mediated diseases. In rheumatoid arthritis patients treated with anti-TNF therapy, higher type I interferon gene expression was associated with lower clinical response rates^41^.

The therapeutic targeting of interferon pathways in IBD has yielded mixed results. While a small study suggested clinical and histological improvements with subcutaneous interferon-α-2A in UC^42^, a subsequent larger trial with pegylated interferon alpha failed to demonstrate clinincal efficacy except for an observed decrease in CRP concentrations^43^. The lack of clinical efficacy following interferon administration is consistent with our findings, which suggest that targeting interferon pathways at baseline may be more likely to improve treatment response. In keeping with this, in a phase I/II study in CD the humanised anti–interferon-γ antibody fontolizumab was associated with reductions in endoscopic severity and CRP levels at higher doses and was well tolerated^44^. A subsequent phase II trial confirmed good tolerability and showed that patients receiving two doses at days 0 and 28 (4 or 10 mg/kg) exhibited doublling in response rate at day 56 compared with placebo^45^.

As the JAK-STAT pathway is a key downstream mediator of interferon receptor activation, our findings suggest the potential for personalised treatment strategies, particularly the use of JAK inhibitors alone or in combination with vedolizumab for non-responders to vedolizumab. Recent systems biology research supports this approach, highlighting the effective combination of JAK inhibition and vedolizumab^46^. Additionally, clinical reports demonstrate success combining vedolizumab with both anti-TNF agents and JAK inhibitors^47^. However, further *in vitro* studies are essential to elucidate the molecular mechanisms influencing variations in treatment outcomes.

Several aspects of our findings warrant further scrutiny. While the interferon signature emerged as a key predictor, the biological mechanisms underlying this association remain to be fully elucidated. Whether suppressed interferon signalling directly mediates response to vedolizumab, or merely serves as a correlated marker of an immune quiescent state, remains unanswered. Functional studies are needed to clarify these causal relationships. Integrating other immune cell types, such as myeloid cells and gut epithelial cells, may provide a more comprehensive understanding of the local immune environment. In addition, as outlined above, our study has limitations related to sample size and cohort heterogeneity. Response definitions and assessment timepoints (10–12, 14, and 30 weeks) and discretionary week-10 dosing varied across cohorts, introducing label heterogeneity that may influence both discovery and classifier performance estimates. The relatively small number of samples in some analyses, particularly in the single-cell RNA-sequencing subset, limits the statistical power to identify specific T cell subsets responsible for the interferon responses. Finally, while our predictive biomarker model showed promising performance, its clinical applicability requires further validation in larger, prospective cohorts and across diverse populations. Moreover, comparator therapies were not evaluated; therefore, the therapy specificity of the baseline interferon signature was not assessed. Integrating transcriptomic data with clinical, biochemical and genetic factors may enhance its predictive accuracy further.

In conclusion, our findings advance the understanding of molecular predictors of vedolizumab response and highlight the interferon pathway as a key immune axis involved in treatment resistance. It also demonstrates the feasibility of a blood-based gene expression classifier across multiple cohorts. Future studies should focus on validating these biomarkers in larger cohorts, exploring their biological roles, and translating them into routine clinical tools to optimise therapeutic strategies and pave the way for personalised treatment in IBD.

## Figure legends

**Supplementary figure 1.** Overview of transcriptomic data generation across cohorts.

**Supplementary figure 2.** Enrichment of interferon response pathways post-treatment in whole blood transcriptomes of responders relative to non-responders across the Kiel and Stockholm cohorts. Bar length represents the –log□□ adjusted p-value, while color indicates the normalised enrichment score (NES) with blue bars denoting pathways downregulated in responders.

## Table legends

**Supplementary table 1.** Baseline characteristics of patients segregated by response.

**Supplementary table 2.** Post-treatment characteristics of patients segregated by response.

## Guarantor of the article

Dr Sreedhar Subramanian

## Author contributions

Conceptualisation: Sreedhar Subramanian, Christopher Probert, Yasmine El Hajj and Paul A Lyons. Data curation: Sreedhar Subramanian, Rachael Slater, Maria Abreu, Sven Almer, Sofie Haglund, Ahmed N. Hegazy, Neha Mishra, Stefan Schreiber and Phillip Rosenstiel. Data analysis: Yasmine El Hajj, George Tang, Sreedhar Subramanian and Paul A Lyons. Writing original draft: Yasmine El Hajj, Sreedhar Subramanian and Paul A Lyons. Writing – review and editing: Yasmine El Hajj, Rachael Slater, Christopher Probert, George Tang, Maria Abreu, Neha Mishra, Sofie Haglund, Stefan Schreiber, Ahmed N. Hegazy, Sven Almer, Phillip Rosenstiel, Paul A Lyons and Sreedhar Subramanian. All authors have read and approved the final version of the manuscript.

This research was supported by the NIHR Cambridge Biomedical Research Centre (NIHR203312). The views expressed are those of the authors and not necessarily those of the NIHR or the Department of Health and Social Care

