## Supplementary figures and tables for "Downregulated Interferon Signalling in T Cells is Associated with Response to Vedolizumab in Inflammatory Bowel Disease"

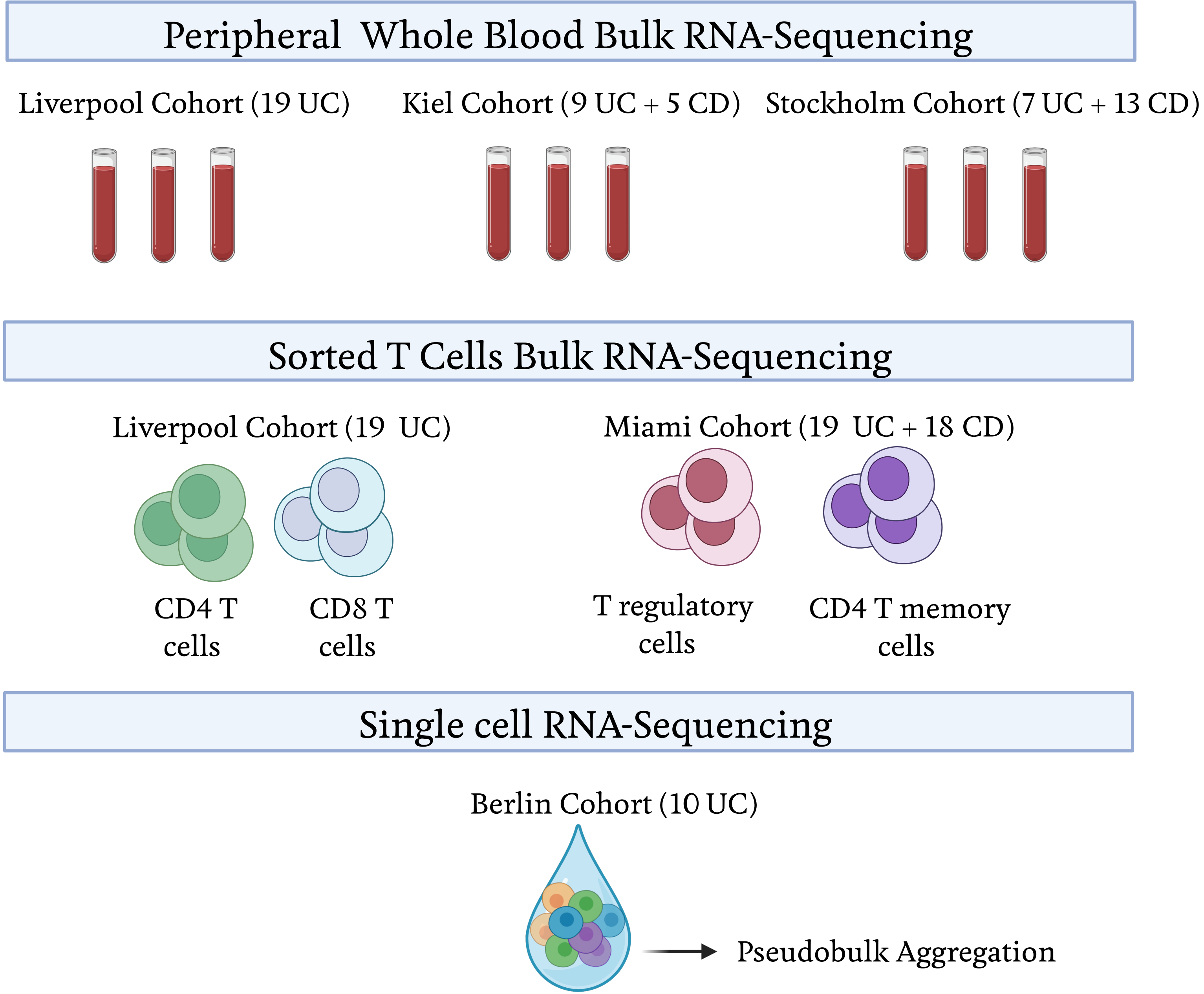
Peripheral Whole Blood Bulk RNA-Sequencing
Liverpool Cohort (19 UC)
Kiel Cohort (9 UC + 5 CD)
Stockholm Cohort (7 UC + 13 CD)
Sorted T Cells Bulk RNA-Sequencing
Liverpool Cohort (19 UC)
Miami Cohort (19 UC + 18 CD)
CD4 T cells
CD8 T cells
T regulatory cells
CD4 T memory cells
Single cell RNA-Sequencing
Berlin Cohort (10 UC)
Pseudobulk Aggregation

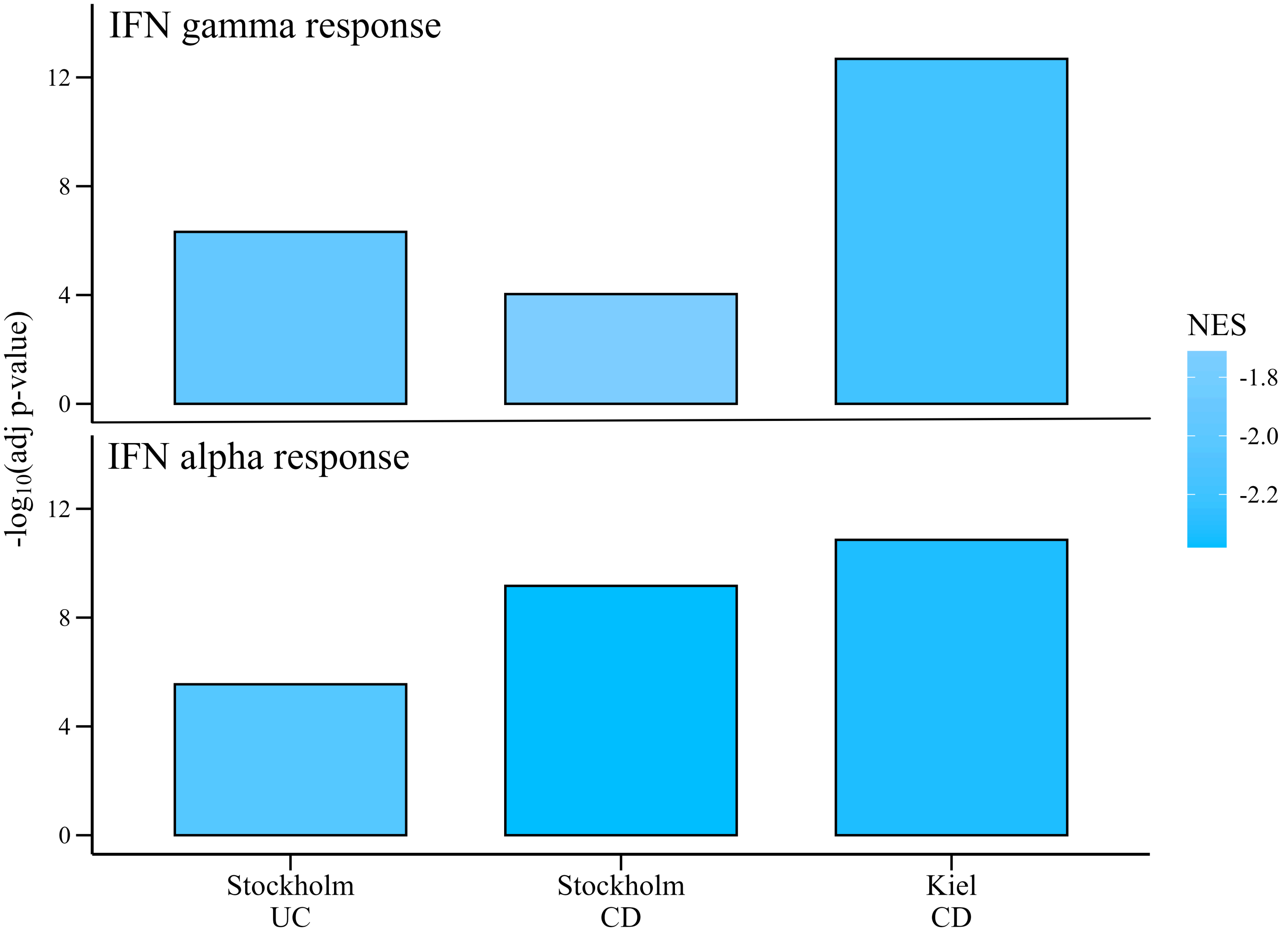

IFN gamma response
IFN alpha response
-log10(adj p-value)
0
4
8
12
Stockholm UC
Stockholm CD
Kiel CD
NES
-1.8
-2.0
-2.2

**Supplementary Table 1.** Baseline characteristics of patients segregated by response

| | Liverpool | | Miami | | Kiel | | Stockholm | |
|---|---|---|---|---|---|---|---|---|
| | **Response n = 15** | **Non-response n = 4** | **Response n = 20** | **Non-response n = 17** | **Response n = 10** | **Non-response n = 4** | **Response n = 10** | **Non-response n = 10** |
| Age (years), mean (SD) | 49 (19) | 54 (22) | 41 (13) | 47 (16) | 41 (17) | 30 (11) | 33 (13) | 40 (17) |
| Sex | | | | | | | | |
| Female, *n* | 5 | 2 | 8 | 11 | 5 | 4 | 3 | 3 |
| Male, *n* | 10 | 2 | 12 | 6 | 5 | 0 | 7 | 7 |
| BMI, mean (SD) | 27 (7) | 29 (5) | 26 (4) | 26 (6) | 27 (5) | 25 (10) | 26 (4) | 24 (3) |
| Disease duration (years), median (IQR) | 7 (10) | 10 (24) | 11 (14) | 16 (16) | 3 (8) | 9 (12) | 10 (13) | 15 (15) |
| Faecal calprotectin (mg/g), median (IQR) | 750 (604), n = 14# | 1111 (1556) | 78 (413), n = 15# | 224 (459), n = 10# | 2005 (684), n = 4# | 345 (850), n = 4# | 568 (1629), n = 7# | 689 (877), n = 9# |
| Partial mayo score, median (IQR) | 5 (4) | 6 (2) | 5 (2) | 3 (1) | 6 (2) | 4 (0) | 6 (2) | 6 (1) |
| HBI score, median (IQR) | *NA* | *NA* | 5 (6) | 5 (2) | 9 (8) | 11 (2) | 5 (1) | 4 (2) |

*#n indicates sample size where data was missing. NA denotes not applicable. – denotes not available.*

**Supplementary Table 2.** Post-treatment characteristics of patients segregated by response

| | Liverpool | | Miami | | Kiel | | Stockholm | |
|---|---|---|---|---|---|---|---|---|
| | **Response n = 15** | **Non-response n = 4** | **Response n = 20** | **Non-response n = 17** | **Response n = 10** | **Non-response n = 4** | **Response n = 10** | **Non-response n= 10** |
| Faecal calprotectin (mg/g), median (IQR) | 750 (604), n = 14# | 1111 (1556) | 86 (129), n = 12# | 216 (700), n = 9# | 291 (452), n = 7# | 360 (315), n = 5# | 345 (1168), n = 6# | 240 (233) |
| Partial mayo score, median (IQR) | 1 (2)* | 5 (3)* | 2 (3) | 4 (3) | 1 (3)* | 5 (1)* | 2 (2)* | 6 (0)* |
| HBI score, median (IQR) | *NA* | *NA* | 2 (3)* | 5 (6)* | 6 (3)* | 11 (0)* | 3 (3) | 4 (2) |

**indicates a signficant difference ($p < 0.05$) between responders and non-responders.*

*#n indicates sample size where data was missing. NA denotes not applicable. . – denotes not available.*
